# A Systematic Review of Multimodal Deep Learning and Machine Learning Fusion Techniques for Prostate Cancer Classification

**DOI:** 10.1101/2025.08.07.25333235

**Authors:** Farhana Manzoor, Vibhuti Gupta, Lubna Pinky, Zhanwei Wang, Zhenbang Chen, Youping Deng, Subash Neupane

## Abstract

Prostate cancer remains one of the most prevalent malignancies and a leading cause of cancer-related deaths among men worldwide. Despite advances in traditional diagnostic methods such as Prostate-specific antigen testing, digital rectal examination, and multiparametric Magnetic resonance imaging, these approaches remain constrained by modality-specific limitations, suboptimal sensitivity and specificity, and reliance on expert interpretation, which may introduce diagnostic inconsistency. Multimodal deep learning and machine learning fusion, which integrates diverse data sources including imaging, clinical, and molecular information, has emerged as a promising strategy to enhance the accuracy of prostate cancer classification. This review aims to outline the current state-of-the-art deep learning and machine learning based fusion techniques for prostate cancer classification, focusing on their implementation, performance, challenges, and clinical applicability. Following the PRISMA guidelines, a total of 131 studies were identified, of which 27 met the inclusion criteria for studies published between 2021 and 2025. Extracted data included input techniques, deep learning architectures, performance metrics, and validation approaches. The majority of the studies used an early fusion approach with convolutional neural networks to integrate the data. Clinical and imaging data were the most commonly used modalities in the reviewed studies for prostate cancer research. Overall, multimodal deep learning and machine learning-based fusion significantly advances prostate cancer classification and outperform unimodal approaches.

## I. INTRODUCTION

### A. CLINICAL BACKGROUND

Prostate cancer (PCa) is one of the leading causes of cancer-related deaths in American men. In 2025, 35,770 deaths of PCa have been projected in the US with 313,780 new PCa cases [1],[2].Incidence rates vary significantly across regions, influenced by differences in screening practices, genetic factors, and healthcare infrastructure. Although early detection through screening has improved survival in high-resource settings, many patients, especially in under-resource settings, still present with advanced disease, highlighting persistent gaps in early diagnosis [3]. Current diagnostic approaches typically start with serum prostate-specific antigen (PSA) testing, digital rectal examination (DRE), biopsy, and Gleason grading. Elevated PSA levels or abnormal DRE findings often prompt systematic transrectal ultrasound-guided biopsies, which carry risks of sampling error, bleeding, and infection. Multiparametric magnetic resonance imaging (mpMRI) has emerged as a valuable non-invasive tool to improve lesion localization and guide targeted biopsies. The Prostate Imaging–Reporting and Data System (PI-RADS) standardizes mpMRI interpretation, yet its diagnostic accuracy remains reader-dependent, with reported sensitivity between 70% and 85% and specificity from 60% to 80% [4].

With the advent of big data collection for precision medicine, a wealth of high-dimensional biomedical data from different modalities is available in routine clinical practice, capturing underlying complex interactions among biological processes. Understanding these interactions is critical while making predictions about complex diseases such as cancer. Artificial intelligence and machine learning techniques have the ability to analyze and integrate vast amounts of biomedical data, enabling the development of comprehensive and personalized solutions that enhance cancer diagnosis, prognosis, and understanding of treatment effects.

### B. RATIONALE FOR MULTIMODAL AI

Single modality data might not be consistent and sufficient to capture the heterogeneity and variability of complex diseases such as cancer to tailor medical care and improve personalized medicine. Combining multiple modalities provides a wealth of complementary and harmonious information that can be exploited for a comprehensive understanding of complex genotype-phenotype associations, better stratify patient populations, and provide individualized care. Deep learning (DL) and machine learning (ML) models trained on single data types—such as imaging alone, histopathology, or genomic profiles—have yielded encouraging results but are constrained by the limited scope of each modality. Imaging-based models may miss molecular heterogeneity, while genomic signatures lack spatial context; clinical variables like PSA kinetics and patient age further modulate disease risk but are often underutilized when used in isolation [5], [6]. These limitations can cause overfitting on small datasets and impede generalizability to broader patient populations.

Integrating heterogeneous data sources—such as imaging, clinical metrics, and molecular profiles—within unified ML/DL frameworks enables the leveraging of complementary strengths: imaging captures spatial morphology, clinical data provides individualized risk context, and molecular profiles uncover underlying biological mechanisms. Such holistic models have demonstrated improved area under the curve (AUC) and better calibration compared to unimodal baselines [7].In PCa, many unimodal approaches were proposed in the past, specifically using histopathological images to detect PCa and classify high vs low risk tumors in the images [8]–[11].However, few of the approaches have leveraged the combined strength of genomics, clinical, and imaging data for PCa risk classification. Thus, this review provides a brief overview of the recent state-of-the-art works done in the multimodal AI, identifies the strengths and limitations and proposes recommendations to address them.

### C. MULTIMODAL DATA TYPES FOR PROSTATE CANCER DIAGNOSIS

1. **Clinical Data**: The clinical data collected for the diagnosis of PCa consists of PSA levels, DRE results, demographic factors, family history, prostate volume, and biopsy results. These metrics provide baseline risk stratification critical for early detection and prognosis, often guiding the initial screening and diagnostic pathways. Clinical nomograms incorporating PSA kinetics and DRE outcomes significantly enhance risk assessment compared to using PSA alone [12], [13].
2. **Genomics Data**: The genomics data collected for the PCa diagnosis consists of germline genetic data, somatic tumor genomic data, and transcriptomics data. Genomic methods assess genetic mutations, gene expression profiles, and epigenetic markers to capture tumor biology at a molecular level. Common genomic techniques include next-generation sequencing (NGS), RNA-seq, methylation profiling, and microarray analyses. Genomic signatures like Decipher, Prolaris, and Oncotype DX provide valuable prognostic information, predicting disease progression and informing personalized treatment strategies [14], [15].
3. **Imaging Data**: Imaging modalities, particularly mpMRI, play a pivotal role in PCa detection, localization, and staging. mpMRI typically includes T2-weighted imaging (T2w), diffusion-weighted imaging (DWI), apparent diffusion coefficient (ADC) mapping, and dynamic contrast-enhanced imaging (DCE). The PI-RADS standardizes interpretation, significantly improving lesion detection accuracy [16], [17]. Additionally, transrectal ultrasound (TRUS) and positron emission tomography/computed tomography (PET/CT) using prostate-specific membrane antigen (PSMA) tracers further refine localization and staging, particularly beneficial in advanced disease states [18], [19].
4. **Molecular Data**: Molecular approaches, including proteomics and metabolomics, evaluate protein expression and metabolite profiles, reflecting tumor metabolism and signaling pathways. PCa metabolomics commonly employs techniques like mass spectrometry and nuclear magnetic resonance spectroscopy to detect biomarkers such as sarcosine, citrate, and choline. These molecular markers significantly improve diagnostic precision and can distinguish aggressive tumors from indolent ones [20], [21].

### D. SCOPE AND OBJECTIVES

In this review, we define multimodal fusion as the systematic integration of diverse modalities-such as imaging, genomics, and clinical data—or multiple data types within a single modality (e.g., MRI, histopathology, CT scans, PSMA PET/CT within imaging)—into a unified deep neural network or ensemble pipeline for PCa classification. We categorize fusion strategies into three major approaches: early (feature-level), intermediate (model-level), and late (decision-level), and examine their implementation in recent studies. The key questions addressed include:

1. What fusion strategies have been explored for PCa classification, and how are “early,” “intermediate,” and “late” fusion defined and implemented in deep-learning pipelines?
2. Which data modalities are being combined—and why? Specifically, beyond mpMRI, what clinical (e.g., PSA, age) or molecular (e.g., gene expression) inputs are integrated, and what preprocessing or harmonization steps are applied?
3. How do different fusion approaches compare in diagnostic performance? What improvements (e.g., AUC, sensitivity, specificity) are reported relative to uni-modal baselines or expert radiologist assessments?
4. What architectural designs and learning frameworks best facilitate multimodal integration, particularly in terms of network backbones (CNNs, transformers, graph models, autoencoders) and attention mechanisms?
5. What are the primary technical and clinical challenges—such as data heterogeneity, missing modalities, overfitting, interpretability, and clinical workflow integration—and how have studies addressed these issues?
6. Where do gaps remain, and what future directions are promising? Which modalities or fusion paradigms are underexplored, and how might emerging methods (e.g., federated learning, explainable AI) advance the field?

To address the above research questions, this review aims to outline the current state-of-the-art multimodal ML/DL fusion techniques for prostate cancer classification, highlight best practices, and identify opportunities for future research. The review is organized according to the following sections: Section II describes the methodology used to retrieve and extract articles for the systematic review, and Section III presents the most common fusion strategies used for multimodal data integration for prostate cancer. Section IV reviews the major data modalities used and the preprocessing approaches used to process the data. Section V focuses on major deep learning architectures used for fusing multimodal data. Sections VI focus on the evaluation metrics used and the comparative analysis of performance across fusion strategies. Section VII provides an overview of benchmark datasets used along with their data sources and section VIII discusses the comparative performance across fusion strategies. Section IX discusses the major findings of the reviewed articles, including current challenges and future research directions. Finally, section Xconcludes the paper.

## II. MATERIALS AND METHODS

### A. SEARCH STRATEGY

We conducted a systematic review of English-language articles using the PubMed, adhering to the Preferred Reporting Items for Systematic Reviews and Meta-analyses (PRISMA) guidelines [22]. The workflow diagram for the systematic identification of scientific literature is shown in Figure 1.

**FIGURE 1.**
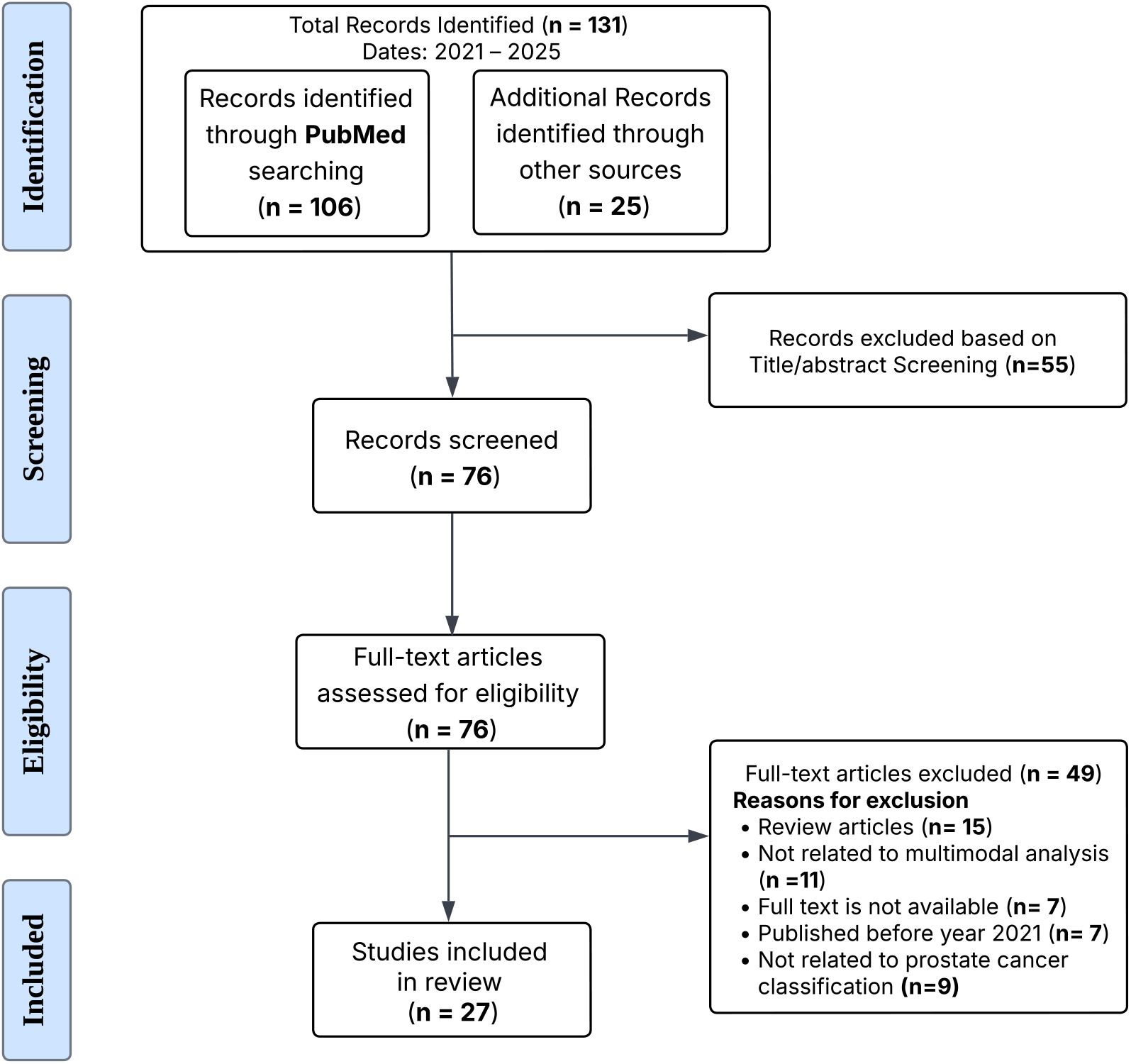
PRISMA workflow for systematic identification of scientific literature. PRISMA flowchart showing the systematic review process for identifying relevant studies on multimodal ML/DL fusion techniques for prostate cancer classification. The diagram illustrates records identified, screened, excluded (with reasons), and included in the final analysis.

**FIGURE 2.**
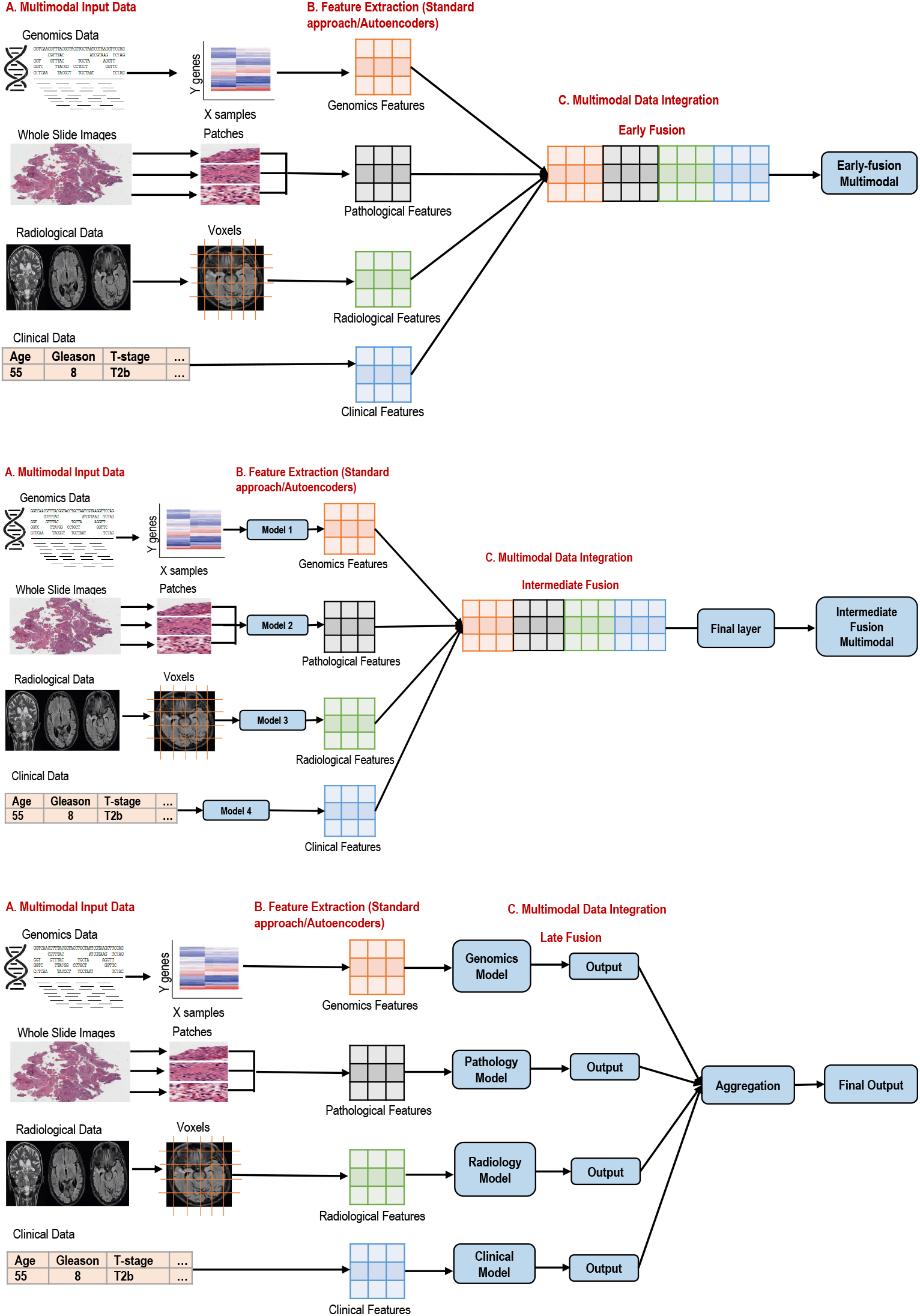
Overview of Fusion techniques

The search terms included various combinations of keywords related to ‘‘Prostate Cancer’’, ‘‘Multimodal’’, ‘‘Deep learning’’, and ‘‘Fusion’’ connected using Boolean operators ‘‘OR’’ (to combine terms within the same domain) and ‘‘AND’’ (to link terms from different domains). Along with the PubMed search, we also retrieved articles by accessing the references of previously published articles on multimodal fusion approaches for PCa classification. We limited our search to review original research articles published, from 2021 to 2025, to ensure we captured the latest advancements in the field.

### B. STUDY SELECTION

Based on the initial search results, we identified 131 studies. These 131 studies were further examined for title/abstract screening. In the screening phase, the title and abstracts of the resulting studies were screened to identify the studies related to multimodal deep learning fusion techniques for PCa classification. This resulted in 76 eligible studies for full text review, as shown in Figure 1 (eligibility section). After identifying the 76 eligible studies for full text review, we applied additional inclusion and exclusion criteria to select the primary studies for our review (details are provided in Figure 1). Studies were eligible if they fulfilled the following inclusion criteria in our review: (1) focused on multimodal analysis; (2) written and published in English; (3) published between 2021 and 2025; (4) full text available rather than abstracts; (5) original studies published in peer-reviewed journals; (6) focused on PCa classification.

Studies were not eligible if they fulfilled the following exclusion criteria in our review: (1) review articles rather than primary research; (2) developed multimodal fusion approaches for other types of cancer; (3) full text not available; (4) published before 2021; (5) focused on other outcomes such as understanding treatment effects, drugs analysis, biomarker identification etc. except PCa classification; (6) using homogeneous data modality for PCa classification. Finally, after applying the inclusion/exclusion criteria to the 76 studies, we identified 27 studies to be included in the detailed review, as shown in Figure 1 in the included phase.

### C. DATA EXTRACTION AND EVALUATION

With the 27 articles identified, we conducted a more detailed review of the shortlisted articles.The data were extracted from all studies meeting our inclusion criteria for the review. It consists of tables containing study information (e.g., authors’ name,year of study), data sources (e.g., hospital collected or public databases) with the number of samples, data modalities, fusion strategies (e.g., early/intermediate/late fusion), evaluation performance, and ML/DL algorithm used (Table 1). In addition, by extracting and synthesizing this wealth of data, we aim to provide a comprehensive overview of the current landscape of multimodal ML/DL fusion approaches for PCa classification. Through this process, we aim to highlight the most widely used approaches, emphasize their advantages, address the challenges,and identify potential areas of improvement. The data for all studies were independently extracted by all authors (VG, FM, LP, ZC, YD, and ZW), and any discrepancies were resolved through mutual discussion among all the authors. The extracted data were finally evaluated by all the authors independently, and consensus was reached through mutual agreement.

**TABLE 1:**
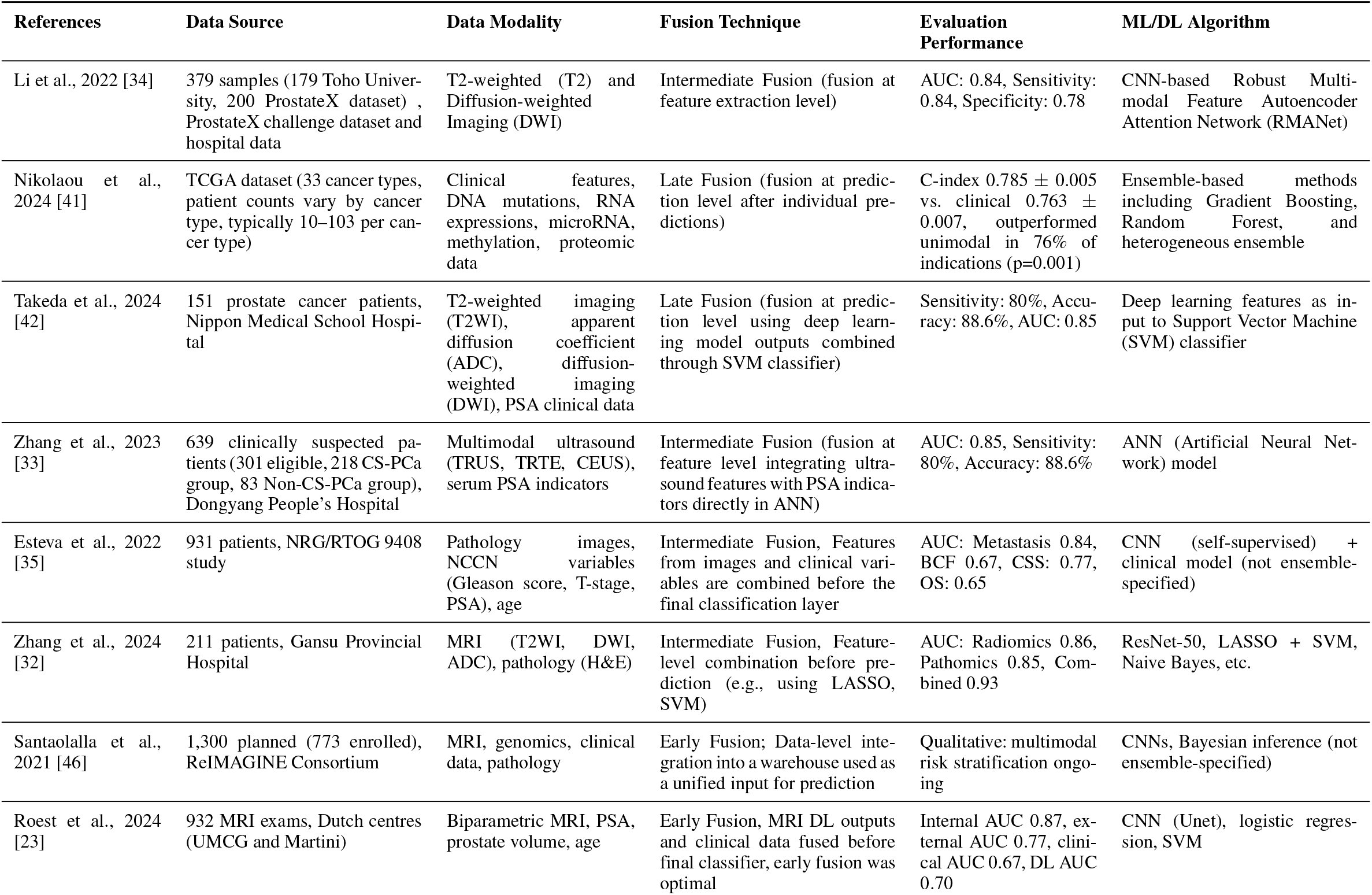
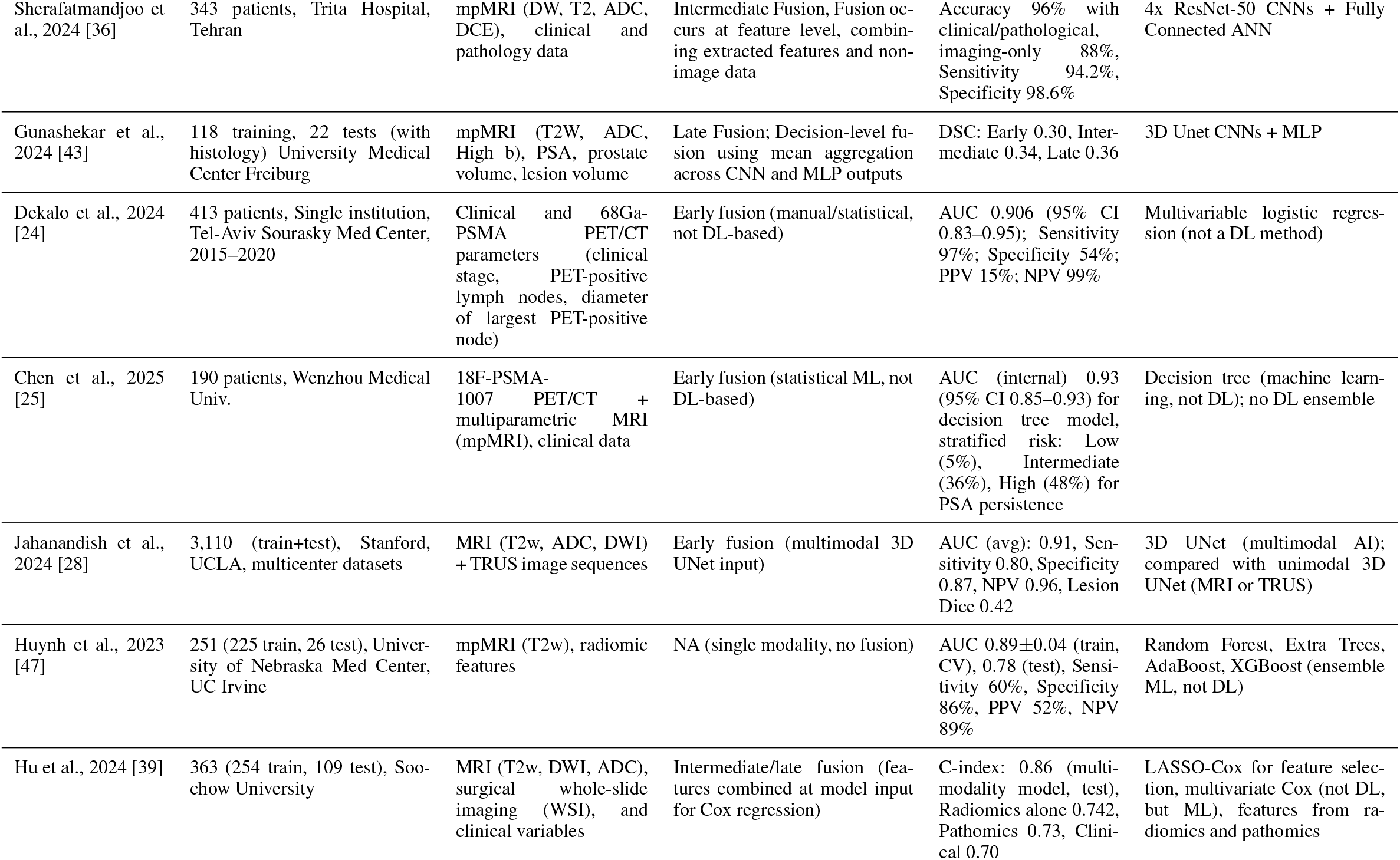
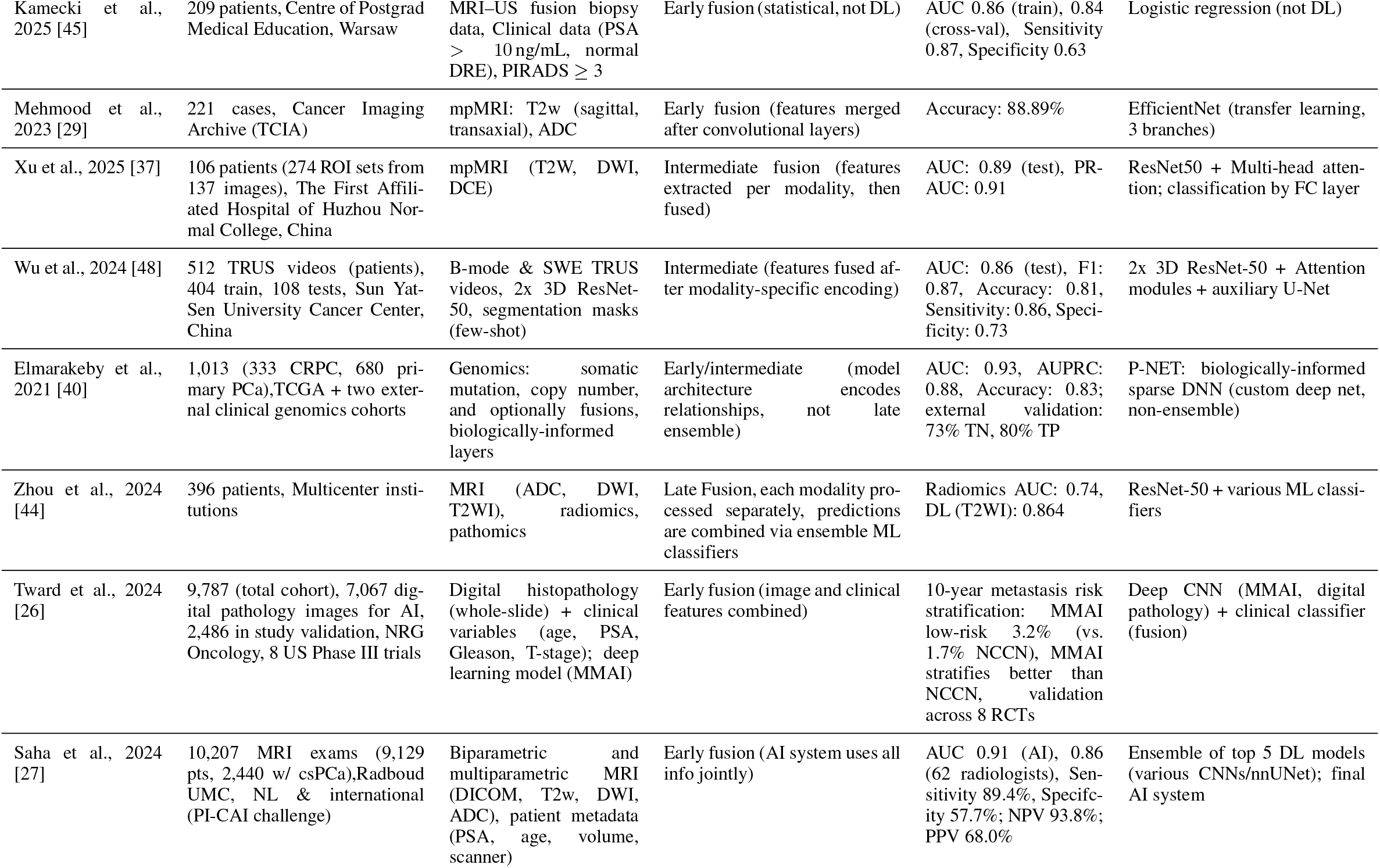
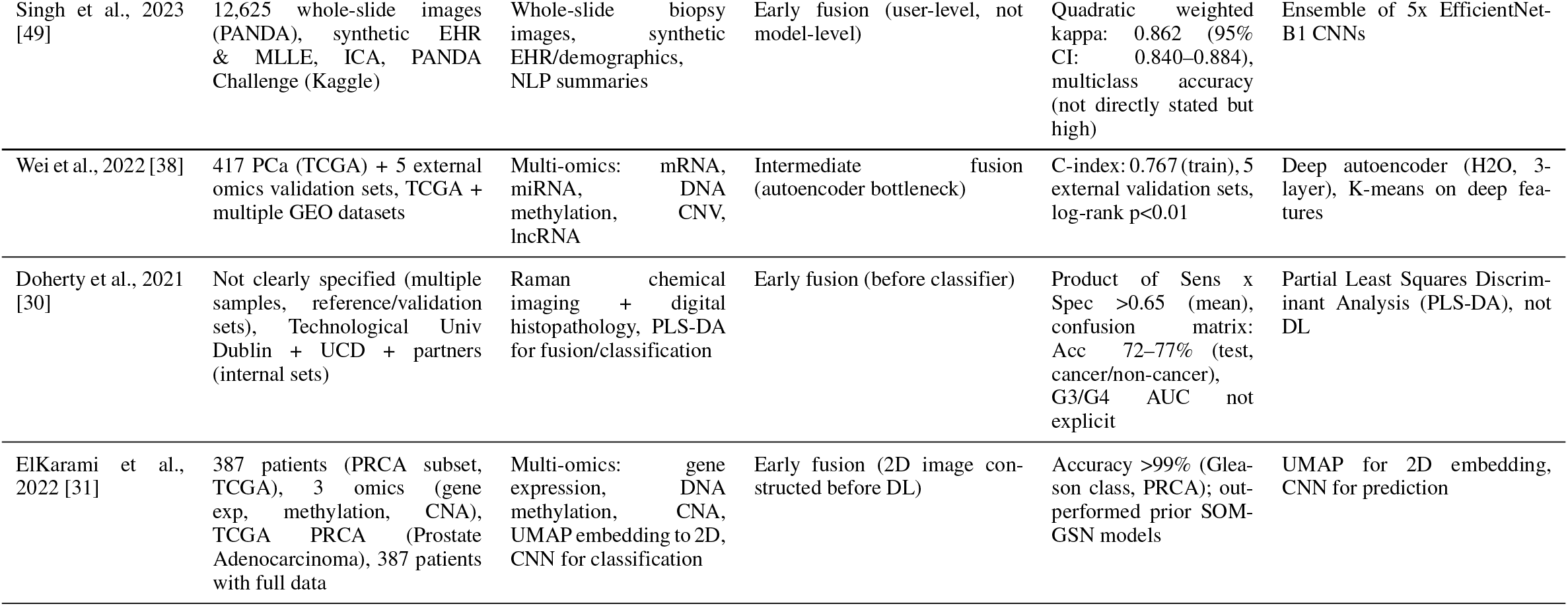
A brief summary of reviewed studies.

## III. TAXONOMY OF FUSION STRATEGIES

### A. EARLY (FEATURE-LEVEL) FUSION

Early fusion involves combining features from all modalities at the initial stage (feature extraction) of the model, enabling joint representation learning from the outset. Each data source—whether T2-weighted and ADC MRI, sequencing data, ultrasound maps, histopathology data, radiomic descriptors, or clinical indices—passes through its own feature extractor (e.g., CNN backbones, texture pipelines, embedding layers), and the resulting vectors are concatenated into one comprehensive representation. This unified feature map is then fed into a downstream classifier. In PCa, the studies [23]– concatenated the clinical variables (i.e., PSA, prostate volume, lesion volume, DRE, gleason, T-stage etc.) with the imaging variables (i.e., MRI, mpMRI, pathology, PSMA-PET/CT) using early fusion. Some of the studies [28]– [30] combined different data types of imaging modalities (i.e.,mpMRI, pathology, PSMA-PET/CT, raman chemical imaging etc.) while one study [31] combined the multiomics data using early fusion. Early fusion excels at learning crossmodal patterns from the outset, but the concatenated feature space can become very high-dimensional, necessitating careful dimensionality reduction or feature selection to mitigate overfitting and manage computational demands [29],[32],[33].

### B. INTERMEDIATE (MODEL-LEVEL) FUSION

Intermediate fusion balances modality-specific learning with joint integration by combining features at a deeper layer within the network. In this paradigm, each modality feeds into its own encoder—separate CNN streams for different MRI sequences, transformer blocks for histopathology, or feed-forward layers for molecular data—and their latent embeddings are merged in shared fusion modules. Attention mechanisms are commonly employed to weight each modality’s contribution dynamically. In PCa, Li et. al [34] combined the T2-weighted and DWI images using intermediate fusion at the feature extraction level while Zhang et. al [33] fused the multimodal ultrasound features (i.e., TRUS, transrectal real-time elastography (TRTE), and transrectal contrast enhanced ultrasound (TR-CEUS)) with the PSA indicators using artificial neural network (ANN). Esteva et. al [35] combined the pathology images with the clinical variables by a joint learning of self-supervised CNN model for images and a catboost model to fuse the clinical features with imaging while Sherafatmandjoo et. al [36] combined the mpMRI images with the clinical and pathology data by extracting image features using ResNet-50 and combining them with the non-image data to train a fully connected ANN. Xu et. al [37]combined the multimodal mpMRI imaging data using intermediate fusion by combining the extracted features using ResNet50 with multi-head attention. The multi-head attention mechanism computes the attention weights and generates weighted feature representations. The combined features were fed into a fully connected layer of neural network to train the model and classify. Wei et. al [38] used intermediate fusion to combine the multi-omics data using autoencoders. Intermediate fusion often outperforms early fusion when ample training data are available, since it allows each modality to develop specialized representations before combining them [17].Nevertheless, these architectures are more complex to train, requiring careful hyperparameter tuning, larger datasets, and sometimes transfer-learning initialization to ensure stable convergence [39], [40].

### C. LATE (DECISION-LEVEL) FUSION

Late fusion maintains independent predictors for each modality and aggregates their outputs at the decision stage. Each unimodal model—such as a CNN on mpMRI, a nomogram on clinical variables, or a survival network on genomic features—is trained separately. Their prediction scores are then combined via ensemble methods like weighted voting, stacking, or meta-learners. In PCa, Nikolaou et. al and Takeda et. al [41], [42] fused clinical variables with genomics and MRI images using late fusion. Nikolaou et. al used an ensemble of gradient boosting, random forest, and other algorithms to train the model; however Takeda et. al used deep learning to extract imaging feature and combine them with clinical features to train a support vector machine (SVM) model. Gunashekar et. al [43] employed decision-level fusion by aggregating outputs from a CNN and a Multi-Layer Perceptron (MLP) using mean pooling, integrating mpMRI data with PSA levels and prostate volume. In contrast, Zhou et al. [44] combined MRI data with clinical information using late fusion and trained ensemble classifiers. Late fusion’s modularity allows easy addition or removal of modalities and gracefully handles missing inputs. Its drawback is that it overlooks deep cross-modal interactions, since modalities are never jointly represented; optimal weighting schemes often rely on heuristic or grid-search approaches [26], [44].

### D. HYBRID AND CASCADED APPROACHES

Hybrid fusion architectures combine early, intermediate, and late strategies within a single pipeline to leverage their respective strengths. A typical cascaded design might begin with early concatenation of imaging and clinical features to generate preliminary risk scores, pass these through intermediate attention or shared layers for refined representation learning, and conclude with a late-stage ensemble that incorporates additional variables such as PSA density or Gleason grade. For instance, one study fused early feature vectors from MRI and ultrasound in a LR nomogram, then refined predictions via a decision-level threshold calibration, achieving AUCs exceeding 0.90 [45]. Other works implement Bayesian latent-class models that blend early modality-specific scores with late survival ensembles to capture both feature-level and decision-level synergies [46]. While hybrid approaches often deliver the highest accuracy, they bring added implementation complexity and reproducibility challenges unless accompanied by clear, containerized workflows and detailed reporting [27], [35].

## IV. DATA MODALITIES AND PRE-PROCESSING

Multimodal artificial-intelligence (AI) systems for PCa classification rely on a diverse array of data sources, each demanding meticulous curation before meaningful fusion can occur. This section reviews different data modalities used for PCa classification and best practices for preparing imaging, clinical, and molecular inputs and harmonizing them across centers and platforms.

### A. IMAGING DATA

#### 1) Multiparametric MRI (mpMRI)

Most multimodal pipelines begin with mpMRI, typically comprising axial or sagittal T2w volumes, DWI with high b-values (≥ 800–1500 s/mm^2^) and corresponding ADC maps; many studies also include DCE sequences to visualise vascular permeability [37]. Raw volumes are first resampled to an isotropic grid——1 mm^3^ in radiomics studies and down to 0.5 mm^2^ for voxel-level segmentation—to equalise spatial resolution across scanners [28]. Intensity standardisation then aligns gray-level distributions: Z-score or Nyúl histogram matching is applied to T2w and DWI, while min–max scaling to the [0, 1] range preserves the biophysical meaning of ADC and DCE maps [34], [47]. Low-frequency bias fields introduced by phased-array coils are commonly removed with N4 correction, especially in older 1.5-T data [39].

#### 2) Cross-modal registration

Spatial alignment is essential when multiple imaging modalities are fused. Voxel-wise fusion models register MRI into the 3-D coordinate system of intra-procedural ultrasound using affine or transformer-based deformable grids [28]. PET/CT is rigidly aligned to mpMRI by maximising mutual information before standardised-uptake-value (SUV) maps are concatenated as additional channels [25]. Whole-slide histopathology images (WSI), in contrast, are colour-normalised and down-sampled to 0.5 *µ*m^−1^; spatial correspondence with MRI is achieved via patient identifiers rather than direct image-to-image transforms [39], [44].

#### 3) Ultrasound and PET extensions

TRUS provides echogenic and stiffness cues that complement MRI. Early fusion approaches simply append TRUS scores to radiomic vectors [42], whereas recent attention networks co-encode 3-D B-mode and shear-wave elastography, refining segmentation with prototype correction [48].^18^For ^68^Ga-PSMA PET adds functional information: SUVmax, total-lesion uptake, and parametric PSMA maps are decay-corrected, Gaussian-smoothed, and incorporated as quantitative channels to predict lymph-node invasion or biochemical persistence [24], [25]. Standardized resampling, normalization, and registration ensure that deep networks learn true pathophysiology rather than protocol artifacts, underpinning reproducible cross-center deployment.

### B. CLINICAL DATA

Core clinical covariates—serum PSA, PSA density (PSAD), age, DRE findings, prostate volume, and Gleason grade—contributearly prognostic context that imaging lacks. Continuous variables are log-transformed and Z-standardised within each cohort before concatenation with radiomic features [23],[45].In repositories containing hundreds of phenotypes (e.g., the ReIMAGINE warehouse), median or multiple-imputation fills sporadic gaps, followed by Spearman or LASSO filtering to retain the most predictive attributes [41], [46]. Deep architectures increasingly embed each scalar into a low-dimensional dense vector so that numerically disparate inputs share a common representational space. In a four-stream mpMRI–clinical–histology network, PSA, age, and prostate volume pass through fully connected layers to create 32-dimensional embeddings that align with 2-D feature maps from ResNet backbones [36]. Saliency analysis shows PSA and PSAD dominate the logits when lesions are small, confirming their complementary weighting.

Encoding strategies vary with model granularity. Nomogram-based logistic-regression models treat age and PSA as continuous covariates but dichotomise DRE (normal versus suspicious) or 5-*α*-reductase-inhibitor use to maintain interpretability [26], [45]. Gradient-boosted trees (CatBoost, XG-Boost) automatically one-hot-encode Gleason grade groups and include missingness indicators, offering native handling of non-linear interactions without explicit scaling (29).Rigorous normalisation and flexible embeddings allow clinical variables to synergise with high-level imaging features, frequently raising the area under the ROC curve (AUC) by 0.07–0.20 over radiology-only baselines [23], [42].

### C. MOLECULAR AND GENOMICS DATA

Although fewer PCa fusion studies incorporate omics, those that do illustrate challenges posed by thousands of heterogeneous molecular features. A pan-cancer survival study downloads mutation, RNA-seq, microRNA, methylation, and proteomics matrices from TCGA, performs median imputation for sporadic missing probes, and prunes to the top 25 univariate features per modality before late-fusion ensembling [41]. This filter discards over 99 % of variables, reducing noise and sparsity.

Biologically informed dimensionality reduction is gaining traction. A pathway-constrained neural network aggregates somatic mutations and copy-number alterations at Reactome pathway nodes, then trains a sparse multilayer network whose topology encodes gene–pathway hierarchies [40]. Weight sharing within pathways limits parameters and retains interpretability without resorting to principal-component analysis.

Beyond dimensionality reduction, embedding-based fusion strategies have emerged as powerful tools for multiomics integration. ElKarami et al. [31] demonstrated that applying UMAP to gene expression data to create a gene similarity network (GSN), followed by RGB encoding of multi-omics features into two-dimensional (2D) images, enables a deep learning pipeline that achieves near-perfect accuracy (>99%) for Gleason grade prediction in prostate cancer. This mid-level (embedding-based) fusion outperformed concatenation and self-organizing map (SOM)-based approaches, suggesting embedding techniques may offer a more biologically meaningful framework for multi-omics data [31]. Wei et al. [38] used a deep autoencoder to integrate five omics layers—messenger RNA (mRNA), microRNA (miRNA), DNA methylation, CNA, and long non-coding RNA (lncRNA)—for relapse risk prediction in prostate cancer, showing robustness via external validation across multiple Gene Expression Omnibus (GEO) datasets. Their framework leveraged bottleneck features from the autoencoder, filtered by least absolute shrinkage and selection operator (LASSO), and clustered by K-means, leading to significantly improved concordance index (C-index) and log-rank statistics, particularly in high-risk patient subgroups. Notably, biological analysis highlighted immune cell infiltration and specific chromosomal instabilities (chromosomes 7 and 8) as key contributors to poor outcomes [38].

Large prospective efforts such as ReIMAGINE collected paired blood, urine, and tissue for germline SNPs, cell-free DNA, and transcriptomics. Preliminary plans include variational-autoencoder compression that maps all omics and clinical covariates into a common 128-dimensional latent space [46]. Across these examples, principal-component analysis, autoencoders, and biologically driven sparsity tame the curse of dimensionality, ensuring that molecular signatures inform rather than overwhelm the classifier.

### D. DATA PREPROCESSING APPROACHES

#### 1) Data Harmonization

Bringing disparate modalities together demands harmonization at both the sample and cohort level.

1. **Handling missing modalities**: Early-fusion convolutional networks simply replace absent channels with zeros, whereas attention transformers drop missing branches and renormalise weights during training—a strategy that maintains AUC when DCE or PET is unavailable [29], [37]. Tabular fusion frameworks impute absent PSA or gene features with cohort medians or include flag variables so tree models can learn patterns of missingness [41].
2. **Batch-effect correction**: Between cohorts, scanner vendors, staining protocols, and sequencing platforms introduce systematic shifts. Histogram matching of T2w and DWI intensities [47]and bias-field correction [39]reduce MRI variance, while ComBat harmonisation is planned for ReIMAGINE’s RNA-seq counts [46]. Domain-adaptation networks map features learned on 3-T scanners to 1.5-T distributions via adversarial losses; combined with rigid + affine MRI-to-TRUS registration, this yields robust cross-platform performance in a Stanford–UCLA multimodal model [28].
3. **Quality control and transparency**:Automated pipelines flag artefacts and track data drift: ReIMAGINE reports a 0.52 % error rate after quality control of more than 40 000 biosamples [46], while nnU-Net-based segmentation discards outliers whose masks deviate more than two standard deviations from the mean prostate volume [23].Publicly released code and benchmark subsets, such as PICAI [27]and PSMA-PET decision tree datasets [24],facilitate external replication, although proprietary data still predominate. Robust imputation, batch effect correction, cross-modal registration, and transparent reporting are therefore indispensable for translating multimodal AI from single-institution prototypes to trustworthy and generalizable clinical tools.

## V. DEEP LEARNING ARCHITECTURES FOR FUSION

### A. CONVOLUTIONAL NEURAL NETWORKS (CNN)

Convolutional Neural Networks (CNNs), especially in 2D and 3D forms, are extensively utilized for processing imaging data in prostate cancer classification tasks. CNN-based fusion techniques generally employ imaging modalities like multiparametric MRI, which includes T2-weighted imaging, diffusion-weighted imaging (DWI), and apparent diffusion coefficient (ADC) maps [28],[29],[34],[42],[47].For instance, a dual-branch CNN combined with self-attention mechanisms and UNet autoencoder features effectively integrates complementary imaging information, enhancing model accuracy [34]. Typically, CNNs utilize fully connected layers at later stages to merge diverse modality-specific features, significantly boosting performance in clinical predictions [29], [42]. CNN architectures have consistently delivered promising diagnostic results, making them valuable tools in clinical settings despite challenges like data variability and segmentation discrepancies [28], [47].

### B. TRANSFORMER AND ATTENTION-BASED MODELS

Transformer and attention-based models have become influential in multimodal fusion due to their ability to identify and leverage intricate relationships between different data types. Cross-modal attention blocks within these models highlight key features across modalities, substantially increasing prediction accuracy. For example, employing multi-head attention mechanisms with various MRI sequences (such as T2-weighted, DWI, and dynamic contrast-enhanced MRI) significantly improves the ability to capture essential diagnostic information [37]. Similarly, SwinTransformer architectures have demonstrated efficacy in aligning MRI and ultrasound data, surpassing both traditional CNN methods and expert radiologist evaluations in lesion detection tasks [28]. The attention mechanisms in these models also enhance interpretability, providing clinicians with more transparent and actionable insights.

### C. GRAPH NEURAL NETWORKS (GNN)

Graph Neural Networks (GNNs) are adept at modeling relationships among data points, which makes them particularly promising for multimodal fusion in medical imaging. Though their application in PCa classification remains limited, analogous methods involving biologically informed neural networks that leverage hierarchical pathway data have shown exceptional predictive accuracy and interpretability [40]. In these approaches, graph aggregators effectively integrate and summarize features from individual nodes (representing lesions or distinct modalities) into a coherent predictive framework. Although specific GNN-based PCa studies are sparse, their demonstrated strengths in relational data modeling suggest a valuable direction for future research.

### D. AUTOENCODER AND VARIATIONAL APPROACHES

Autoencoder-based methods, particularly those employing variational frameworks, have been effective in creating unified representations of heterogeneous data types. One notable example involves a dual-branch autoencoder model integrating T2w MRI and ADC maps. This approach extracts modality-specific features and improves classification performance significantly over single-modality methods [34]. Autoencoder architectures facilitate the reduction of high-dimensional data to more manageable latent spaces while retaining essential diagnostic information. Although variational approaches have potential advantages in quantifying uncertainty within latent spaces, explicit applications in prostate cancer diagnostics remain limited and represent an exciting avenue for further exploration. The deep autoencoder-based framework from Wei et al. [38]represents one of the most robust omics fusion strategies to date, successfully integrating up to five distinct omics layers for relapse risk stratification in prostate cancer. External validation across five GEO cohorts supported the generalizability of this approach, underscoring the potential of autoencoder bottleneck representations as universal latent spaces for complex, high-dimensional omics data. Embedding-based strategies, as shown by ElKarami et al. [31],further expanded this landscape by encoding multi-omics relationships into 2D RGB images via UMAP, enabling CNN classifiers to leverage both local and global structural information for high-accuracy prediction.

### E. ENSEMBLE AND META-LEARNING TECHNIQUES

Ensemble and meta-learning methods have emerged as robust strategies for effectively integrating diverse modality predictions, offering enhanced performance and reliability through uncertainty estimation. These methods typically involve stacking or blending predictions from individual modality-specific models. For instance, late fusion techniques, which combine modality-specific risk scores such as clinical, imaging, and molecular data, have consistently demonstrated improvements in predictive capabilities across cancer diagnostics, including PCa scenarios [41]. Meta-learning ensemble methods integrating predictions from radiomics, clinical data, and pathology have similarly shown enhanced accuracy and robustness [32], [39], [44]. Techniques involving prototype-based feature refinement, combined with attention mechanisms in ultrasound imaging modalities, further enhance specificity and interpretability [48]. These ensemble methods not only increase predictive performance but also effectively quantify uncertainties, providing critical support for clinical decision-making.

In summary, various deep learning architectures—CNNs, transformer and attention-based models, GNNs, autoencoders, and ensemble/meta-learning methods—each offer unique advantages for multimodal data fusion in PCa classification. The reviewed studies illustrate substantial benefits achievable through the integration of different data sources, underscoring promising avenues for future methodological developments and enhanced clinical applications.

## VI. EVALUATION METRICS AND STATISTICAL ANALYSIS

### A. CLASSIFICATION METRICS

In PCa classification tasks involving multimodal learning, the evaluation typically includes metrics such as AUC-ROC, accuracy, sensitivity, specificity, and the F1-score. The AUC-ROC metric is frequently utilized because it effectively summarizes model performance across various classification thresholds. For instance, a CNN-based model that integrated MRI and ADC imaging achieved strong performance with an AUC of 0.84, sensitivity of 0.84, and specificity of 0.78, highlighting its effectiveness in distinguishing clinically significant prostate cancer from benign cases [34]. Ensemble approaches that combine clinical, imaging, and molecular data also demonstrated robust performance, with high concordance indices outperforming single-modality models [41]. Accuracy, sensitivity, and specificity offer complementary insights and are particularly critical for clinical decision-making. For example, a multimodal model combining MRI and transrectal ultrasound (TRUS) showed good sensitivity (0.80) and specificity (0.87), underscoring its clinical potential for accurate prostate lesion detection [28]. The F1-score, which balances precision and recall, is particularly useful for datasets with class imbalance. An EfficientNet-based model integrating multiparametric MRI demonstrated a high F1-score of 89.47%, reflecting balanced predictive performance across different cancer severity classes [29].

### B. CALIBRATION AND CLINICAL UTILITY

Calibration curves and decision curve analyses (DCA) play a crucial role in evaluating the clinical usefulness of predictive models. Calibration curves assess how closely the predicted probabilities match observed outcomes, providing clinicians confidence in model predictions. Models utilizing radiomics and pathomics data have shown strong calibration, meaning their predicted outcomes align well with actual clinical outcomes, thus enhancing trust and utility in clinical practice [32], [39]. Decision curve analysis quantifies the clinical net benefits of predictive models across a range of decision thresholds. Models integrating clinical, imaging, and histopathological information have consistently demonstrated superior net benefits compared to traditional clinical nomograms. These multimodal approaches have practical implications for clinical decision-making, particularly in risk stratification and treatment decisions, highlighting their real-world utility [39].

### C. SIGNIFICANCE TESTING

Statistical significance testing is critical for validating the comparative performance of predictive models. DeLong’s test is commonly employed to compare AUC-ROC curves between models, providing robust statistical evidence of performance differences. For instance, DeLong’s test confirmed that multimodal models significantly outperform single-modality models (clinical-only or imaging-only), reinforcing the statistical reliability of integrating multiple modalities [23], [42].

Evaluation of embedding-based multi-omics fusion models has yielded near-perfect accuracy, precision, recall, F1-score, and AUC in large public cohorts, with the UMAP+CNN pipeline from ElKarami et al. [31] reporting >99% accuracy and AUC 0.99 for Gleason score prediction, and Wei et al. [38] demonstrating robust C-index (>0.75) and significant survival stratification in multiple external sets. Beyond deep learning models, machine learning-based fusion of label-free Raman chemical imaging with digital histopathology, as demonstrated by Doherty et al. [30], achieved reasonable accuracy (confusion matrix accuracy approximately 72–77%) and clinically relevant sensitivity/specificity combinations in PCa classification, illustrating the breadth of multimodal strategies currently under exploration. Bootstrap confidence intervals offer another robust method for assessing performance reliability, accounting for variability due to sampling. Repeated cross-validation combined with bootstrap methods have consistently demonstrated reliable ROC-AUC results, particularly in radiomic models predicting PCa recurrence, thereby ensuring confidence in the generalizability of model predictions across diverse patient groups [47].

### D. EXPLAINABILITY METRICS

Interpretability and explainability have become increasingly important in medical AI to improve transparency and clinician acceptance. SHapley Additive exPlanations (SHAP) [50]values offer detailed insights into model predictions by quantifying the contribution of individual features. For instance, biologically informed neural networks have utilized SHAP analyses effectively, revealing significant genomic pathways and genetic alterations relevant to PCa progression, thus improving model interpretability and facilitating clinical understanding [31].

Gradient-weighted Class Activation Mapping (Grad-CAM) [51] is another essential visualization tool, particularly beneficial for imaging-based models. Grad-CAM visualizations have been effectively utilized in multimodal MRI and TRUS models, clearly indicating which regions of input images significantly influence the model’s decisions. This has substantially enhanced the interpretability of model predictions, making these tools more useful in clinical decision-making, especially for targeted biopsy strategies [44], [48].

In summary, the evaluation metrics and statistical methodologies used in multimodal PCa classification collectively cover aspects of performance evaluation, clinical relevance, statistical rigor, and interpretability. Metrics such as AUC-ROC, accuracy, sensitivity, specificity, and F1-score provide comprehensive assessments of predictive performance. Calibration and decision curve analyses further confirm the practical clinical value of predictive models. Significance testing methods like DeLong’s test and bootstrap confidence intervals ensure that observed performance improvements are statistically robust. Finally, explainability tools such as SHAP and Grad-CAM enhance transparency and trust, facilitating better clinical integration and acceptance of multimodal deep learning models. These comprehensive evaluation approaches ensure that multimodal models not only demonstrate superior predictive capabilities but are also practically applicable, statistically validated, and inherently interpretable, thus promoting their effective adoption in clinical practice.

## VII. BENCHMARK DATASETS AND EXPERIMENTAL PROTOCOLS

Multimodal PCa fusion studies draw on a blend of open and institutional cohorts to capture diverse patient populations and imaging/omics modalities. Public resources such as PROSTATEx [52] (T2-weighted, DWI/ADC and clinical labels) and The Cancer Genome Atlas (TCGA) [53](multiomics with matching demographics and outcomes) underpin many investigations [29], [34], [41]. In parallel, several groups have released single-center or multicenter MRI and pathology collections—e.g., institutional mpMRI from Freiburg and Tehran [36], [43], NRG Oncology trial data with histopathology and long-term follow-up [26], [35], and hybrid MR-TRUS biopsy sets from Stanford and UCLA [28]. Recent multi-omics fusion studies have leveraged The Cancer Genome Atlas Prostate Adenocarcinoma (TCGA-PRAD) datasets and multiple GEO validation cohorts to ensure robustness and generalizability. For example, both Wei et al. and ElKarami et al. systematically trained and validated their multimodal models on publicly available omics data, demonstrating the importance of external cohorts for credible performance benchmarking [31], [38]. To ensure robust performance estimates, authors employ a mix of internal cross-validation, held-out splits, and fully external test sets. Small to mid-sized single-site studies typically report five- or ten-fold cross-validation results, reserving 10–20% of cases for an unseen hold-out fold [39], [44]. By contrast, larger or multicenter efforts often use one institution as an external validation cohort—training on combined sites and testing entirely on the held-out center—to better gauge generalizability [23], [28]. Several prospective analyses also adopt time-based splits, training on earlier cases and evaluating on more recent scans, to simulate real-world deployment [42].

Preprocessing pipelines share three core steps. First, automated or semi-automated segmentation delineates prostate anatomy and lesions—nnU-Net variants are the de facto standard in many MRI studies [23], [28]. Second, intra- and intermodal registration aligns images (e.g., rigid registration between T2 and ADC) and, when applicable, maps MRI to ultrasound or histology space [28]. Third, feature extraction transforms raw inputs into quantitative descriptors: classical radiomic features via established toolkits [44], deep-learning embeddings from pretrained backbones for pathology and ultrasound [32], [48], and handcrafted clinical indices such as PSA density and Gleason score.

### A. REPRODUCIBILITY CONSIDERATIONS

Although most studies describe cohorts, splitting schemes, and preprocessing steps, full reproducibility often remains aspirational. A subset of authors share code repositories, Dockerfiles, and model weights, enabling direct replication of key experiments [23], [40], [48]. However, many rely on protected institutional datasets without public release, and method descriptions sometimes lack parameter details or seed declarations [23], [24], [32]–[36], [39], [42], [44], [47]. To foster transparent benchmarking and accelerate clinical translation, the community would benefit from broader adoption of open-access data portals, containerized workflows, and adherence to reporting guidelines such as TRIPOD-AI for prognostic modeling.

## VIII. COMPARATIVE PERFORMANCE ACROSS FUSION STRATEGIES

### A. EARLY VS. INTERMEDIATE VS. LATE FUSION

The timing of multimodal data fusion significantly influences PCa classification performance. Early fusion strategies typically integrate raw or minimally processed features from different modalities at the input stage. For example, the nnUNet-based early fusion model integrating MRI and TRUS achieved a robust performance, yielding an ROC-AUC of 0.91 and sensitivity of 0.80, demonstrating the benefit of immediate cross-modality interactions [28]. Similarly, early fusion of clinical parameters with deep-learning-based MRI suspicion scores has consistently shown superior diagnostic accuracy, particularly in distinguishing clinically significant PCa [18].Intermediate fusion models integrate modality-specific features at hidden layers, leveraging learned representations. A notable intermediate fusion example employed an EfficientNet architecture to fuse multiparametric MRI modalities, achieving a high accuracy of 88.89% and F1-score of 89.47%, clearly outperforming unimodal approaches [29].

Recent embedding-based integration methods, such as UMAP-driven RGB image fusion of multi-omics data, have yielded state-of-the-art accuracy in Gleason grading [31](>99%) and have outperformed previous SOM-based methods. Likewise, autoencoder-based fusion of five omics layers [38]enabled external validation across multiple GEO cohorts, underscoring the advantage of mid-level, learned representations for robust classification and risk stratification. In parallel, fusion of non-imaging modalities such as Raman spectroscopy and histopathology images, using partial least squares discriminant analysis (PLS-DA), has been piloted as a non-deep learning strategy for multimodal diagnostics in PCa, demonstrating promise for non-invasive, label-free detection but with lower accuracy than deep learning-based approaches [30].

Late fusion models integrate modality-specific predictions at the decision level, typically via ensemble or meta-learning techniques. A late fusion strategy combining radiomics, pathomics, and clinical features using a Cox regression-based nomogram demonstrated substantial predictive performance (C-index of 0.86), emphasizing the utility of integrating predictions derived independently from each modality [39]. Additionally, a late-fusion ensemble approach across various genomic, clinical, and imaging modalities demonstrated significant performance improvements over single modalities, highlighting the robustness of decision-level integration [41].

### B. MODALITY CONTRIBUTION ANALYSIS

Ablation studies provide critical insights into modality contributions, delineating the incremental value each modality adds to overall model performance. MRI modalities consistently emerge as critical contributors. Ablation analyses indicate significant performance deterioration when ADC or diffusion-weighted imaging modalities are excluded, underscoring their pivotal role [29], [34]. Similarly, incorporating TRUS data alongside MRI substantially improves lesion detection accuracy, demonstrsxssating valuable spatial and textural contributions [28]. Ultrasound dsemodalities, particularly shear-wave elastography (SWE), significantly enhance predictive accuracy, confirming their utility in multimodal approaches [48]. Clinical data also plays an essential role, as evident by marked performance reductions when parameters such as PSA levels and prostate volume are excluded [42], [45].

## IX. DISCUSSION

### A. PRINCIPLE FINDINGS

Across 27 studies integrating imaging, clinical, molecular, and pathological data for prostate cancer assessment, multimodal fusion consistently outperforms single-modality models. Early feature-level fusion is the most common strategy (12/27) as shown in supplementary figure S3, leveraging complementary information before classification, and frequently yields AUCs in the 0.85–0.93 range. Intermediate fusion—often employing attention or concatenation of learned representations—demonstrates robust discriminative performance (AUC 0.86–0.91) while modeling cross-modal interactions more flexibly. Late decision-level fusion is less prevalent but provides interpretability through ensemble voting or nomograms, with AUCs typically 0.78–0.88. Importantly, large multicentre and trial datasets [26]–[28], [35] show that rigorous external validation and standardized pipelines bolster generalizability and calibration.

Analysis of the comprehensive comparison table indicates that intermediate fusion strategies generally exhibit the most consistent and robust performance across multiple metrics, particularly accuracy, sensitivity, specificity, and F1-score. This consistency can be attributed to intermediate fusion’s capability to exploit learned modality-specific representations effectively, thus maximizing complementary information and minimizing modality-specific noise or redundancy. Although early fusion methods have shown promising results, particularly in leveraging raw feature interactions at early stages, their performance can be variable depending on preprocessing rigor and data quality. Late fusion provides flexibility and robustness by integrating independently validated predictions, especially beneficial when dealing with heterogeneous or sparse datasets. However, intermediate fusion emerges as the optimal balance, providing a more universally effective framework for robust multimodal integration in PCa classification.

Most of the reviewed studies used clinical and imaging data combined (13/27) followed by imaging data of various types (MRI, CT, Pathology) (9) and multi-omics (3) as shown in Supplementary figure S2. There are only 1 study conducted using all clinical, imaging, and genomics data for PCa classification research and 1 with clinical and genomic data. This shows that the research on multimodal fusion using all types of data is still in a growing stage. In terms of ML/DL techniques employed in the studies, most of the studies used CNNs, followed by ensemble learning techniques, as shown in Supplementary Table S1. Many studies used conventional machine learning algorithms (i.e., LR, tree-based and Bayesian algorithms, discriminant analysis and least absolute shrinkage and selection operator) too along with the deep learning techniques. This shows that the deep learning architectures and ensemble-based machine learning frameworks are widely used for multimodal integration in PCa studies.

### B. CHALLENGES AND RECOMMENDATIONS

#### 1) Data Heterogeneity and Scarcity

Multimodal deep learning fusion strategies for PCa classification consistently face challenges related to data heterogeneity and scarcity. The diversity of imaging modalities—such as mpMRI, ultrasound, and pathology images—introduces variability that complicates standardized preprocessing and harmonization across datasets [28], [34]. Additionally, many reviewed studies were constrained by limited dataset sizes, reducing the generalizability of the developed models and increasing the likelihood of biased results [29], [32], [34]. Similar challenges have been observed in other cancer types, emphasizing the necessity of standardized multimodal datasets and international collaborations [54].Future research must prioritize cross-center collaborations and standardized data collection protocols to enhance model robustness and ensure broader clinical applicability.

#### 2) Overfitting in High-Dimensional Spaces

High-dimensional data integration, typical in early and intermediate fusion methods, often leads to overfitting, particularly when sample sizes are small or insufficiently diverse. Techniques such as dimensionality reduction, rigorous feature selection via methods like LASSO regression, and transfer learning are essential to mitigate overfitting [29],[32], [41], [47]. Approaches such as regularization, ensemble methods, and deep learning optimization techniques have shown promise in reducing overfitting in other biomedical contexts [55]. Further exploration into these techniques could enhance the stability and generalizability of prostate cancer classification models.

#### 3) Interpretability, Clinical Trust, and integration into clinical workflow

Although multimodal deep learning methods demonstrate improved diagnostic accuracy, interpretability remains a critical challenge affecting clinical trust and adoption. Several studies addressed this by implementing attention mechanisms and feature refinement approaches to highlight clinically significant regions [28], [34], [48]. Comparable efforts in breast cancer imaging have demonstrated the value of interpretability frameworks such as Grad-CAM for clinician acceptance and confidence [56]. Enhancing interpretability through explainable AI techniques such as saliency mapping and attention visualization is recommended to build trust and facilitate clinical integration.

It is important to note that embedding-based multi-omics fusion methods such as RGB encoding are currently limited to three omics due to the RGB(Red-Green-Blue encoding) color space constraint [31]. Furthermore, although external validation in [31], [38] supports generalizability, current public omics datasets are modest in size, and the field would benefit from larger multicenter initiatives. Non-deep learning fusion of chemical imaging and digital pathology provides a parallel avenue for multimodal PCa detection, although performance remains inferior to deep learning-based fusion—highlighting the need for continued innovation and cross-modality bench-marking [30].

Integrating multimodal AI tools into existing clinical workflows poses operational challenges. Systems must be robust, easy to use, and seamlessly integrated into current diagnostic pathways without disrupting established practices [23], [42], [45]. Lessons from radiology and oncology indicate that adoption requires thorough validation through prospective clinical trials, comprehensive training for clinical staff, and clear guidelines on interpreting and utilizing AI-generated predictions [57].

#### 4) Ethical and Privacy Concerns

Ethical considerations and patient privacy remain significant concerns, particularly regarding data sharing and the handling of sensitive genomic and imaging information. Several studies discussed data anonymization and ethical approval procedures, yet few comprehensively addressed broader ethical implications or privacy-preserving mechanisms such as federated learning [23], [39], [46]. Wider adoption of federated learning, demonstrated in cardiovascular research, underscores its potential to safeguard privacy while enabling collaborative AI training [58]. Ensuring adherence to data protection standards and clearly defined ethical frameworks is imperative for sustainable advancement and clinical adoption of multimodal AI.

## X. CONCLUSIONS AND FUTURE RESEARCH

We conducted a systematic review of multimodal deep learning and machine learning fusion techniques for PCa classification. In recent years, there has been increasing interest in the application of multimodal AI approaches in PCa classification. The majority of studies used early feature-level fusion with the most consistent and robust performance with intermediate fusion. Most of the studies used clinical and imaging data combined. However, very few studies used all types of modalities including imaging, clinical, and genomics. The primary challenges include data heterogeneity and limited availability across imaging protocols, difficulties in interpretability and clinical integration, as well as ethical and privacy concerns. Our findings indicate that improvements may be achieved through the use of harmonized multicenter datasets, the integration of explainable AI methods and intuitive visualization tools into clinical workflows, rigorously validating multimodal models in diverse, multicenter datasets, and the adoption of federated learning or secure multiparty computation alongside transparent governance frameworks to ensure patient confidentiality. Looking ahead, the field should incorporate emerging data sources—such as liquid biopsies, advanced radiomic features, and wearable biosensors—to enhance predictive modeling and support continuous patient monitoring. Federated and privacy-preserving learning approaches will be crucial for harnessing distributed data while maintaining patient confidentiality, whereas real-time and edge deployments can deliver AI-driven support directly at the point of care, including in interventional settings. To ensure reproducibility, it is essential to establish standardized reporting guidelines, shared benchmark datasets, and openly accessible codebases. Finally, advancing explainable multimodal AI—by integrating attention-based fusion mechanisms with post hoc interpretability frameworks—will be essential to bridging the gap between technical innovation and clinical trust. Such models are critical for fostering clinician confidence, enabling transparent understanding and effective use of AI-generated insights in prostate cancer diagnosis and management.

## Data Availability

All data produced in the present work are contained in the manuscript.

## XI ACKNOWLEDGMENT

This research was funded by the, AIM-AHEAD Coordinating Center, award number OTA-21-017, and was, in part, funded by the National Institutes of Health, United States Agreement No. 1OT2OD032581 and P20GM103466, U54MD007601, U54GM138062 and U54HG013243. The work is solely the responsibility of the authors and does not necessarily represent the official view of AIM-AHEAD or the National Institutes of Health.

## Notes

### Competing Interest Statement

The authors have declared no competing interest.

